# Mislocalization and clearance of neuronal Rhes as a novel hallmark of tauopathies

**DOI:** 10.1101/2020.10.27.20220954

**Authors:** Alexander J. Ehrenberg, Kun Leng, Israel Hernandez, Caroline Lew, William W. Seeley, Salvatore Spina, Bruce Miller, Helmut Heinsen, Martin Kampmann, Kenneth S. Kosik, Lea T. Grinberg

## Abstract

The farnesyltransferase inhibitor lonafarnib reduces tau inclusion burden and atrophy in familial tauopathy models by inhibiting farnesylation on the Ras GTPase, Rhes, and activating autophagy. While hinting at a role of Rhes in tau aggregation, it is unclear how translatable these results are for sporadic forms of tauopathy. We used a combination of quantitative pathology using multiplex immunofluorescence for Rhes, several tau post-translational modifications, and single nucleus RNA sequence analysis to interrogate Rhes presence and distribution in human cortical neurons and Rhes relation to tau and TDP-43 changes. snRNA data suggest that Rhes is found in all cortical neuron subpopulations, not only in striatum cells. Histologic investigation in hippocampal formation from multiple postmortem cases in five different tauopathies and healthy controls and TDP-43 proteinopathy showed that nearly all neurons in control brains display a pattern of diffuse cytoplasmic Rhes positivity. However, in the presence of abnormal tau, but not TDP-43 inclusions, the patterns of neuronal cytoplasmic Rhes tend to present as either punctiform or fully absent. Our findings reinforce the relevance of the link between Rhes changes and tau pathology suggested by *in vivo* and *in vitro* models of tauopathy and support a potential clinical application of lonafarnib to tauopathies.

## INTRODUCTION

Despite being the most frequent underlying cause of dementia, tauopathies remain without effective treatments and are increasing in prevalence [1, 2]. Tauopathies encompass many distinct neuropathological entities that feature the accumulation of abnormally folded tau protein followed by associated neuron and synapse loss. Alzheimer’s disease (AD) is the most common tauopathy and features both 3- and 4-repeat tau inclusions. Among the sporadic tauopathies, progressive supranuclear palsy (PSP), corticobasal degeneration (CBD), and argyrophilic grain disease (AGD) show 4-repeat tau inclusions predominantly while Pick’s disease (PiD) features 3-repeat tau inclusions. Although the overwhelming majority of tauopathies occur sporadically, more than 50 mutations in *MAPT* lead to abnormal tau accumulation and different expression patterns of frontotemporal lobar degeneration in an autosomal dominant pattern of inheritance [3-5]. These mutations are the basis for biomedical research in tauopathies, as they can be incorporated into transgenic cell lines or *in vivo* systems that model tauopathies; however, it is unclear how generalizable findings from these familial models are for the more common sporadic tauopathies.

Immunotherapies have been the most common class of the drugs brought to clinical trials for tauopathies; however, they have, thus far, produced lackluster results [6, 7]. Other approaches have attempted to modulate post-translational modifications of tau [7-10] or reduce MAPT mRNA expression using antisense oligonucleotides [11]. The well-documented dysfunction in autophagy across tauopathies [12, 13] establishes autophagy modulation as an attractive strategy to enhance clearance of aggregated tau across a wide spectrum of tauopathies [13, 14]. These efforts have shown some promise in preclinical settings and may become effective treatment strategies for tauopathies.

Farnesyltransferase inhibitors were initially developed for cancer treatment by acting on farnesyl protein transferase, a necessary catalyst of protein CaaX farnesylation, to inhibit Ras oncoproteins [15]. A pharmacological class effect of farnesyltransferase inhibitors is enhanced autophagy [16]. The farnesyltransferase inhibitor, lonafarnib was successfully used *in vitro* and in mouse models of familial tauopathy to reduce the burden of tau inclusions by enhancing autophagic activity [17]. Specifically, lonafarnib acts through the Ras guanosine triphosphatase, Ras homolog enriched in striatum (Rhes) to activate autophagic degradation of tau aggregates. Rhes has been shown to suppress autophagy by binding to and activating the mammalian target of rapamycin (mTOR) [18]. Through sequestration of Beclin-1 from inhibitory binding to Bcl-2, though, Rhes can also activate autophagy independent of mTOR [19]. When farnesylation is blocked by an inhibitor such as lonafarnib, Rhes dissociates from membranes, thus allowing it to sequester Beclin-1 and upregulate autophagy [17].

Although promising, the translatability of these model-based observations to treating tauopathies, particularly sporadic tauopathies, using a farnesyltransferase inhibitor is unclear. Furthermore, little is known about the pattern of Rhes expression in human neurons, particularly outside the striatum and in the context of neurodegenerative conditions.

Rhes is encoded by the gene *RASD2* and was thought to be predominantly expressed in the striatum [17, 20]. Some studies suggest that Rhes expression in specific neuronal subpopulations is the basis of selective vulnerability to Huntington’s disease, a genetic neurodegenerative condition causing profound striatal and mild cortical neuronal loss [21]. However, tauopathies show only mild striatal involvement, but significant involvement of cortical and other subcortical structures. The evidence for Rhes expression in human cortical neurons is scant with just one study, to our knowledge, showing scattered expression of *RASD2* mRNA throughout the cortices of humans and mouse models using autoradiography [20].

An in-depth understanding of Rhes expression, localization within neurons, and disease-related changes in cortical regions vulnerable to tauopathies, if any, is a necessary step to inform whether lonafarnib should be explored as a treatment for tauopathies. To fill this gap, we used multiplex immunofluorescence and single nucleus RNA sequencing in human brain tissue from healthy controls and a broad range of sporadic tauopathies to characterize and measure phenotype changes in Rhes and its relationship to the accumulation of tau aggregates.

## MATERIALS AND METHODS

### Participants and Neuropathological Assessment

Cases were sourced from the Neurodegenerative Disease Brain Bank at the University of California, San Francisco Memory and Aging Center. Consent for brain donation was obtained from subjects or next of kin following a protocol approved by the Institutional Review Board of the University of California, San Francisco.

Upon autopsy, the brains were slabbed fresh into coronal slabs and either fixed in 4% neutral buffered formalin or frozen within 24 hours of death. Slabs undergoing formalin fixation was transferred to PBS-Azide after 72-hours of fixation and stored at 4°C. A standardized set of 26 tissue blocks from neurodegenerative regions of interest are dissected from these slabs and embedded into paraffin blocks. Immunohistochemical and basic histological stains were used to assess neurodegenerative lesions using antibodies for phospho-Ser202 Tau (CP13, Mouse, 1:250, courtesy of Peter Davies), TDP-43 (Rabbit, 1:2000, 10782-2-AP, Proteintech, Chicago, IL), β-amyloid (Mouse, 1:500, MAB5206, Millipore, Billerica, MA), α-Synuclein (Mouse, 1:5000, LB509, courtesy of J. Trojanowski and V. Lee). A final neuropathologic diagnosis was obtained after consensus (LTG, SS, and WWS) using currently accepted guidelines [22-24].

For the immunohistochemical component of this study, we selected 18 cases (Table 1) encompassing healthy controls (HC) and five sporadic tauopathies (AD, PSP, CBD, PiD, and AGD). Half of the cases were female with a mean (sd) age of death of 72 (10) years. Inclusion criteria included: lack of more than one neuropathological diagnosis, lack of an Axis I psychiatric disorder, lack of non-dementia neurological disorder, lack of gross non-degenerative structural neuropathology, and a postmortem interval below 24 hours. Healthy control cases were free of any clinical record of cognitive decline, neurological, or a non-incidental neuropathological diagnosis.

### Tissue processing, immunohistochemistry, and microscopy

#### Quantitative histopathological analysis

All 18 cases encompassing the HCs and tauopathies were used in this analysis. The paraffin block containing the hippocampal formation at the level of the lateral genicular body was sectioned at 8µm. Sections underwent deparaffinization in xylene then stepwise rehydration from 100% to 80% Ethanol. For antigen retrieval, sections were cycled through an autoclave set at 121°C for five minutes in a 10 solution of 0.1M citrate buffer (pH 6.0) with 0.05% tween-20. The solution was left to cool to room temperature then transferred to 1x phosphate-buffered saline with 0.05% tween (PBST) for five minutes to wash. Sections were then incubated in 5% milk with 0.05% tween for 30 minutes to block non-specific binding. Tissue underwent primary incubation for 18 hours overnight at room temperature with a cocktail of NeuN (266 004, 1:600, Synaptic Systems), Rhes (GTX85428, 1:200, GeneTex), and PHF-1 (phospho-Ser396/Ser404 Tau, 1:600, Gift from Peter Davies) diluted in the 5% milk/tween. In additional sections from AD cases, PHF-1 was replaced with AT100 (phospho-Thr212/Ser214 Tau, 1:800, MN1060, Invitrogen), 1f3c (acetyl-Lys274 Tau, 1:1000, Gift from Li Gan), Tau-C3 (ΔAsp421 Tau, 1:300, AHB0061, Invitrogen), or 1:500 MC1 (Conformational Tau, Gift from Peter Davies).

Sections were washed with PBST and then underwent a 90-minute incubation in 1:400 Goat anti-Mouse IgG (H+L) Alexa Fluor 488 (A-11001, Invitrogen), 1:400 Goat anti-Rabbit IgG (H+L) Alexa Fluor 546 (A-11010, Invitrogen), and 1:400 Goat anti-Guinea Pig IgG (H+L) Alexa Fluor 647 (A-21450, Invitrogen) in PBST. Following washing with PBST, sections went through a two minute wash in 70% ethanol to a 40-minute wash in Sudan black B. Excess Sudan black was removed with two, ten-second washes in 70% ethanol then immediately washed in dH_2_O. Sections were cover-slipped with Prolong with NucBlue (P36981, Invitrogen).

Sections were scanned using a Zeiss Axio Scan.Z1 slide scanner (Molecular Imaging Center, University of California, Berkeley) with a Plan-Apo 10X/0.45 NA objective using DAPI (Zeiss Filter Set 49), eGFP (FS 38 HE), Cy3 (FS 43 HE), and Cy5 (FS 50) filters. We qualitatively examined the intraneuronal signal pattern in the Rhes channel. Based on consensus (AJE and LTG), we identified three different patterns of intraneuronal Rhes deposition. Next, we defined the regions of interest (ROI) for quantitative analysis. We delineated 1×1mm frames in the entorhinal cortex (EC) and cornu ammonis 1 (CA1) sector of the hippocampus with the assistance of an expert neuroanatomist (HH). All neurons in each ROI were counted, categorizing each neuron as PHF-1 positive or negative and as one of the three Rhes subtypes. A similar analysis was conducted with the combinations including AT100, 1f3c, Tau-C3, and MC1; however, sections were scanned with a Plan-Apo 20X/0.8 NA objective and instead of the one 1×1mm counting frame, three 650×650µm regions were delineated from CA1.

#### Analysis of association with TDP-43 proteinopathy

The hippocampal formation of two additional cases of frontotemporal lobar degeneration due to TDP-43 type A (Case 1, 64 year-old female with corticobasal syndrome; Case 2, 66 year-old female with behavioral variant frontotemporal dementia) were examined. After completing deparaffinization and rehydration, endogenous peroxidase was quenched in 3% hydrogen peroxide (H2O2) and 80 % methanol for 30 minutes. Antigen retrieval was conducted in an autoclave set at 121°C for five minutes in a solution of 0.1M citrate buffer (pH 6.0) with 0.05% tween-20. After cooling to room temperature, the sections were washed in PBST.

Sections were incubated in 5% milk with 0.05% tween for 30 minutes then underwent primary antibody incubation overnight at room temperature. The primary antibody combination included anti-phospho-S409/410 TDP 43 (1:200, Rat, TAR5P, gift of Manuela Neumann, Munich, Germany) [25], Rhes (GTX85428, 1:200, GeneTex), and NeuN (266 004, 1:600, Synaptic Systems).

Sections were washed with PBST and then underwent a 90-minute incubation in 1:400 Goat-anti-rat IgG (H+L), HRP conjugate (R-05075-500, Advansta), 1:400 Goat anti-Guinea Pig IgG (H+L) Alexa Fluor 546 (A-11074, Invitrogen), and 1:400 Goat anti-Chicken IgY (H+L) Alexa Fluor 647 (A-32933, Invitrogen) in PBST. The sections underwent a 15-minute tyramide signal amplification with a solution of 1:100 AF488 Tyramide (B40953, Thermo Fisher) and 1:100 of 100x H2O2 in 1x tris buffered saline. Following washing with PBST, the sections went through a 2-minute wash in 70% ethanol to a 40-minute wash in Sudan black B. Excess Sudan black was removed with two, ten-second washes in 70% ethanol then immediately washed in dH2O. The sections were incubated at 1:50,000 in DAPI, washed with PBS, and coverslipped with Prolong (P36980, Invitrogen).

#### Analysis of intracellular distribution

One paraffin block containing the hippocampal formation at the level of the lateral genicular body was sectioned at 8µm for one of the AD cases (59 year-old male). The section underwent deparaffinization in xylene then stepwise rehydration from 100% to 80% ethanol. For antigen retrieval, the section was cycled through an autoclave set at 121°C for five minutes in a solution of 0.1M citrate buffer (pH 6.0) with 0.05% tween-20. The solution was left to cool to room temperature then transferred to 1x phosphate-buffered saline with 0.05% tween (PBST) for five minutes to wash. The section was then incubated in 5% milk with 0.05% tween for 30 minutes to block non-specific binding. Tissue underwent primary incubation for 18 hours overnight at room temperature with a cocktail of 1:600 NeuN (266 004, anti-guinea pig, Synaptic Systems), Rhes (GTX85428, 1:200, anti-rabbit, GeneTex), and MAP-2 (NB300-213, 1:900, Novus) diluted in the 5% milk/tween.

The section was washed with PBST and then underwent a 90-minute incubation in 1:400 Goat anti-Rabbit IgG (H+L) Alexa Fluor 488 (A-11008, Invitrogen), 1:400 Goat anti-Guinea Pig IgG (H+L) Alexa Fluor 546 (A-11074, Invitrogen), and 1:400 Goat anti-Chicken IgY (H+L) Alexa Fluor 647 (A-32933, Invitrogen) in PBST. Following washing with PBST, the section went through a two minute wash in 70% ethanol to a 40-minute wash in Sudan black B. Excess Sudan black was removed with two, ten second washes in 70% ethanol then immediately washed in dH2O. The section was incubated at 1:50,000 in DAPI, washed with PBS, and coverslipped with Prolong (P36980, Invitrogen).

### Gene expression

Processed single-nucleus RNA-sequencing data from Leng et al. [26] was downloaded from Synapse (Synapse ID: syn21788402). This dataset represents single-cell gene expression information and metadata from the EC, labeled as “EC_allCells.rds”, and superior frontal gyrus (Brodmann area 8), labeled as “SFG_allCells.rds”, from postmortem brain samples of HC and individual across the neuropathological spectrum of AD. The log-scaled counts per million (CPM) of *RASD2* transcripts in cells from the cell type subpopulations identified in the EC and SFG of Braak stage 0 cases were used to assess expression patterns. Next, the mean log CPM of *RASD2* transcripts within groups 2 and 4 of excitatory EC neurons [26] was taken for each case (n=10) and compared between Braak stages 0, II, and VI [27] using a one-way ANOVA.

### Statistical analysis

To examine differences in the relative proportions of neuron populations belonging to each phenotype between diagnoses, the proportion of neurons belonging to each phenotype was computed as the number of neurons featuring each of the three phenotypes divided by the total number of neurons counted per case. Wilcoxon-Mann-Whitney tests were used to test the null hypothesis that no significant difference exists in the proportion of total neurons counted that present as Rhes Type-I between healthy controls and each disease in pairwise comparisons.

To test for differences in PHF-1 tau inclusion burden within each neuronal Rhes phenotype population, the proportions of each neuronal Rhes phenotype that were PHF-1 positive and PHF-1 negative were computed for each region of interest (EC and CA1) for each case. Kruskal-Wallis rank-sum tests were used to test the null hypothesis that there was no significant difference in the burden of PHF-1 tau inclusions between the three neuronal Rhes phenotypes in each disease. At a group-level, combining all cases, one-way ANOVA’s were used to test the null hypothesis that there was no significant difference in the burden of PHF-1 tau inclusions between the three neuronal Rhes phenotypes in the EC or CA1. For both Kruskal-Wallis rank-sum tests and one-way ANOVA’s, cases without a computed proportion (i.e., those with a denominator of 0) were omitted from the analysis.

The analyses involving other post-translational modifications of tau did not involve statistical tests as they were exploratory. Instead, the percentage of neurons positive for each post-translational modification classified as each Rhes phenotype was computed and averaged between the two cases.

The alpha level for all analyses was set to 0.05. Statistical analysis was conducted in the software R, and graphs were produced with the R package ggplot2.

## RESULTS

### Rhes is ubiquitously expressed in neurons of healthy controls, in a cytoplasmic granular distribution

Several studies have suggested that Rhes expression is mostly limited to the striatum, thus the name Rhes. Using immunofluorescence, we detected Rhes signal in neurons throughout the allocortex (Figure 1a). In healthy controls (HC), we found Rhes signal in over 99% of EC and CA1 NeuN+ cells. In these neurons, Rhes presents with a cytoplasmic granular distribution extending into the dendrite (Figure 1b-i).

**Figure 1:**
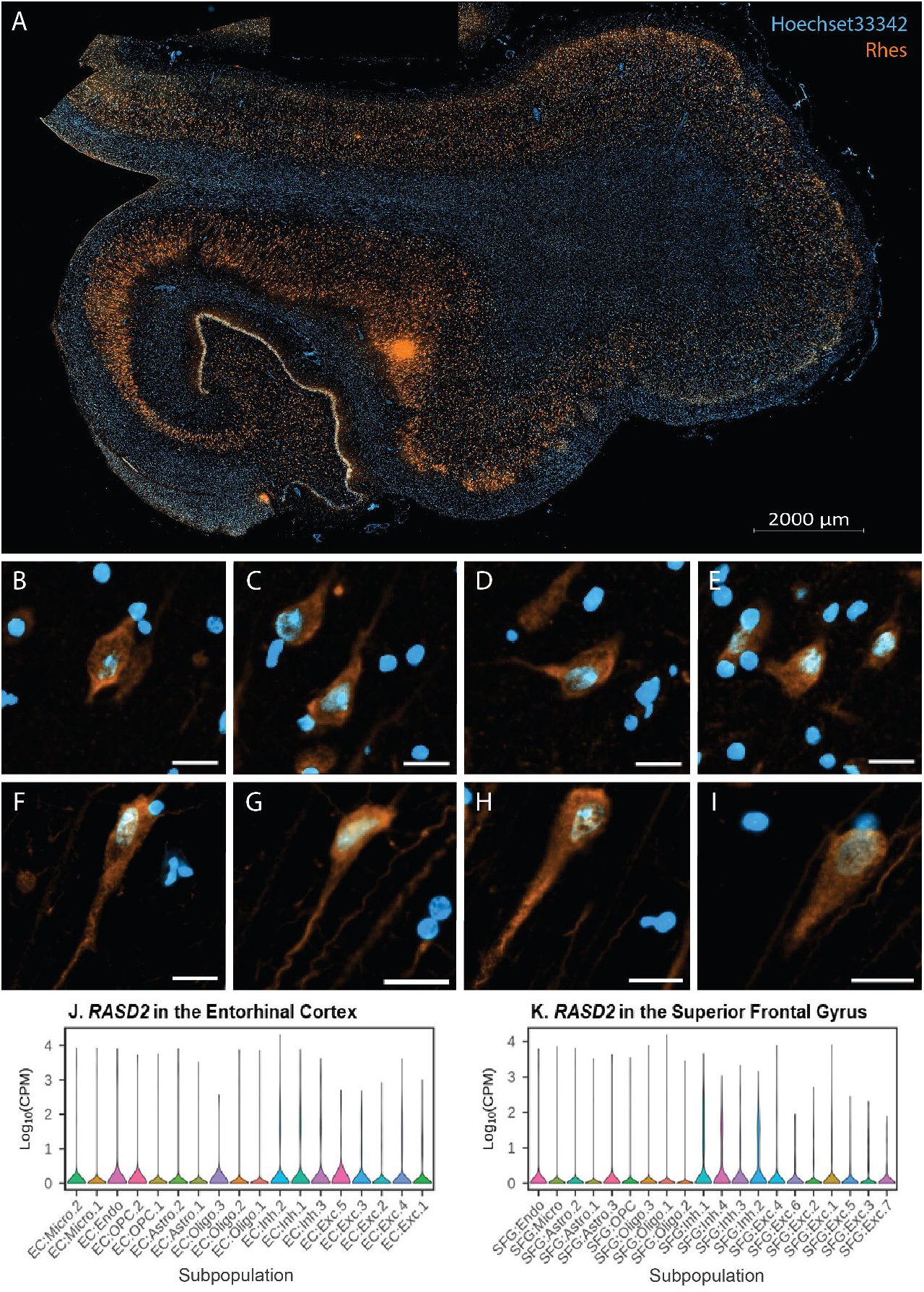
Rhes (orange) is ubiquitously expressed throughout the human hippocampal formation (A). In entorhinal cortex (B-E) and cornu ammonis 1 (F-I) neurons, Rhes has a granular appearance in the cell body (Hoechst 33342 in blue), extending into proximal segments of neuronal processes (Type-I). All scale bars B-I correspond to 20 µm. The corresponding transcript, *RASD2* is found in all excitatory and inhibitory neuronal subpopulations of the entorhinal cortex (J) and superior frontal gyrus (K).

To validate the findings, we analyzed single-nucleus RNA sequencing data from the EC (an allocortex) and superior frontal gyrus (a neocortex) from a different cohort [26]. Within the Braak stage 0 cases (n=3), we detected *RASD2* transcripts in all excitatory and inhibitory neuronal populations of these two regions (Figure 1j-k).

### Neurons of individuals with tauopathies present as three subtypes of Rhes based on intraneuronal localization

We qualitatively examined neurons in the EC and CA1 of all 18 cases (Table 1) representing HC and five tauopathies (AD, PSP, CBD, PiD, and AGD) and defined three distinct phenotypes of Rhes distribution in neurons. Altogether, these three phenotypes accounted for 100% of neurons. Rhes Type’ Diffuse’ neurons featured granular Rhes staining in the cell body extending into the dendrite (Figure 2a). Type ‘Punctiform’ neurons have large cytoplasmic Rhes+ puncta with a variable granular background, extending into the dendrite (Figure 2b). Type ‘Absent’ neurons feature an absence of Rhes staining in the cell body, and variable positivity in the dendrite (Figure 2c).

**Figure 2:**
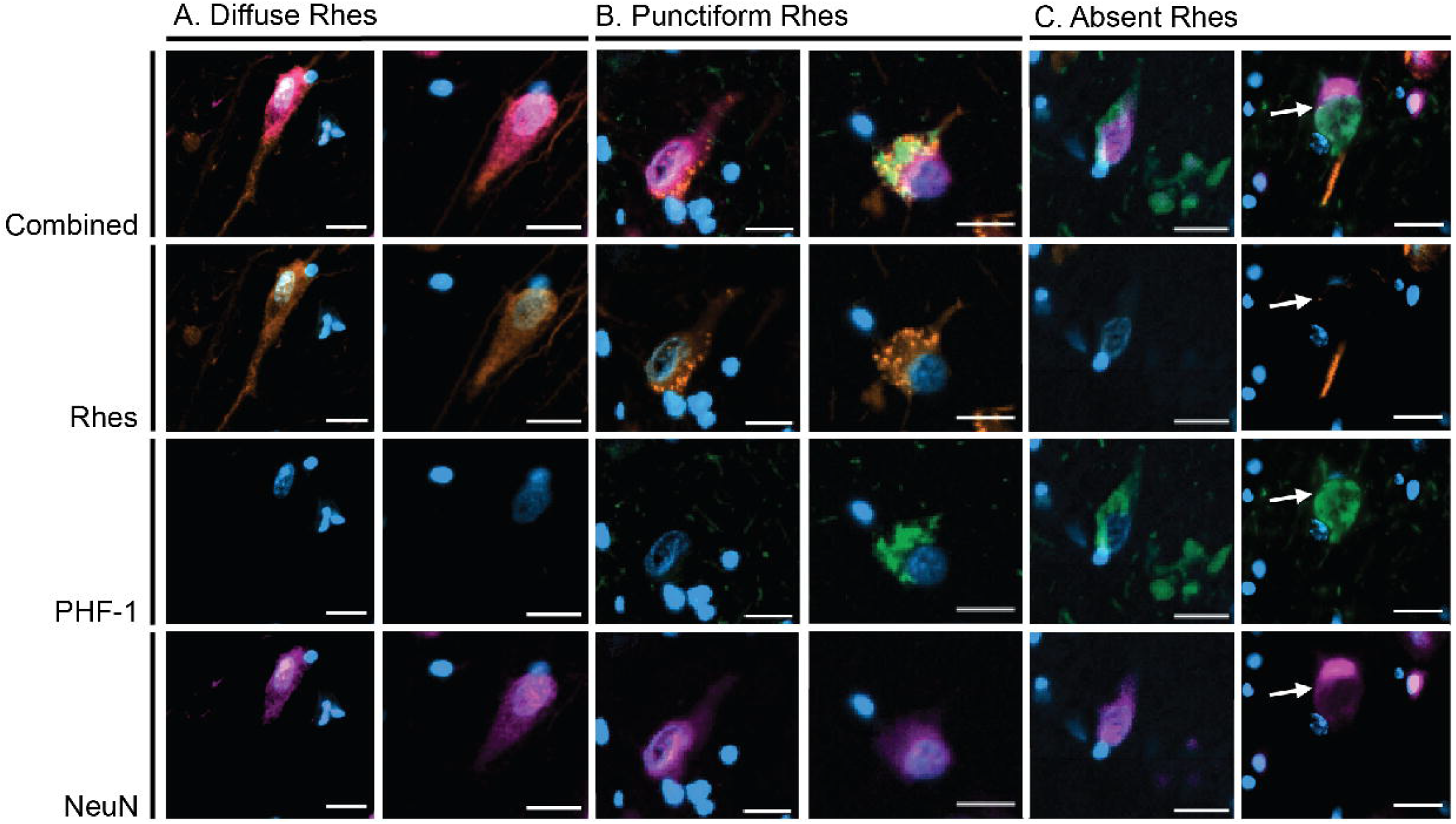
Diffuse Rhes (orange) neurons (A) feature diffuse Rhes staining in the cell body extending into the proximal axonal segment. Punctiform Rhes neurons (B) feature punctiform Rhes signal in the cell body and often co-occur with PHF-1 (pink) positive tau inclusions. Absent Rhes neurons (C) feature clearance of Rhes from the cell body and often co-occur with PHF-1 positive tau inclusions. Blue, DAPI; Orange, Rhes; Green, PHF-1; Pink, NeuN. All scale bars correspond to 20 µm.

To confirm that Rhes is expressed in the dendrites, histological slides of the human hippocampal formation underwent multiplex immunostaining with Rhes, MAP2 and NeuN. We observed that Rhes overlaps with MAP2 signal in the neuronal processes of all three subtypes, indicating that it is present in dendrites. (Figure 3).

**Figure 3:**
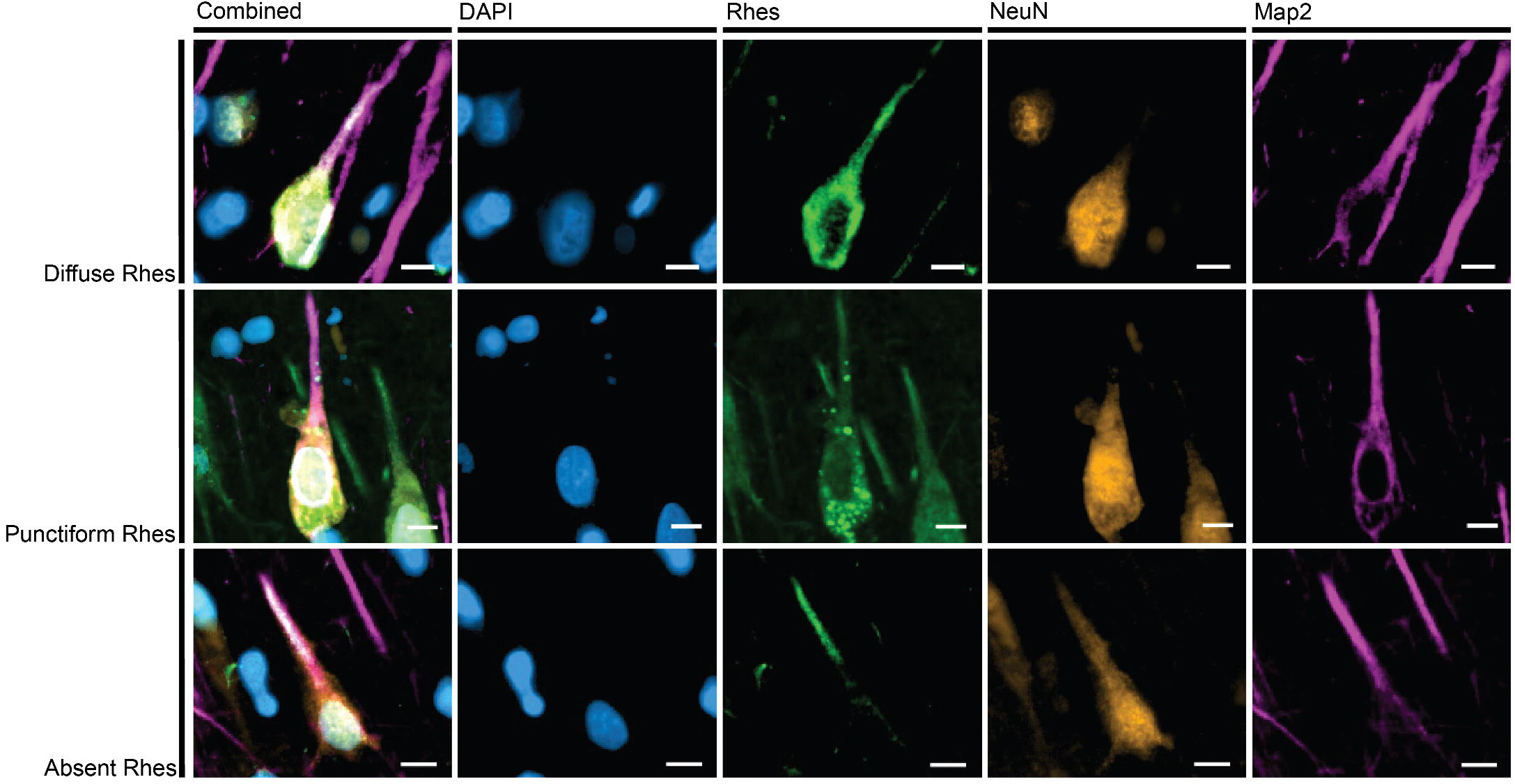
In all three Rhes neuronal presentations, projections of Rhes (green) overlap with Map2 (pink). This indicates that Rhes occurs in dendrites. Examples here from CA1 of a 64-year old female with AD (A3B3C3). Scale bars correspond to 10 µm.

### The relative abundance of Rhes neuronal phenotypes in AD and other tauopathies

In HC, over 99% of neurons in the EC and CA1 showed a diffuse Rhes phenotype, but the Punctiform and Absent phenotypes were frequently seen in the tauopathy groups. A one-way ANOVA showed that the proportion of neurons with a diffuse Rhes phenotype was significantly different between tauopathies in the EC (p = 0.028) and CA1 (p < 0.001). This finding suggested that in tauopathies, intraneuronal Rhes assumes a punctiform presentation or is cleared from the neuronal cytoplasm in affected neurons.

AD, PiD, and PSP showed the highest proportion of neurons with Punctiform or Absent Rhes phenotypes (Figure 4a-b, Table 2). Compared to HC, where 99% of neurons presented with a diffuse Rhes phenotype, only 86% of EC neurons and 50% of CA1 neurons had diffuse Rhes in AD. Similarly, 85% of EC neurons and 66% of CA1 neurons had diffuse Rhes in PiD, and 91% of EC neurons and 89% of CA1 neurons had a diffuse Rhes in PSP. Notably, in AGD, a 4 repeat tauopathy featuring scarce to moderate numbers of cytoplasmic neuronal inclusions in the EC and CA1, the proportion of neurons with diffuse Rhes phenotypes did not show a significant decrease compared to HC. Similarly, CBD, a 4-repeat tauopathy with mild pathology in CA1 and EC, showed no significant differences in the proportion of neurons with Rhes Type-I phenotypes compared to HC.

**Figure 4:**
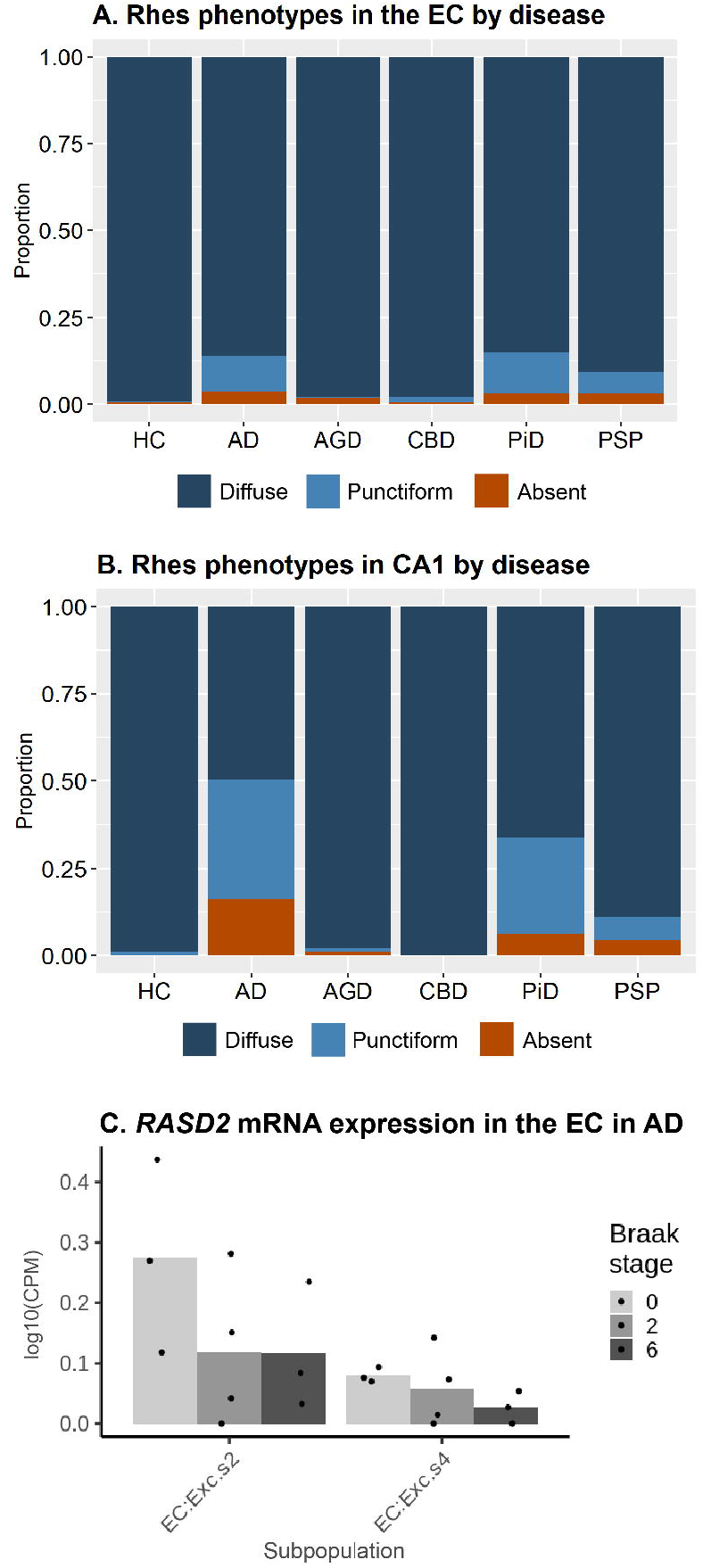
The relative proportions of Rhes intraneuronal phenotypes across tauopathies in the entorhinal cortex (A) and CA 1 (B). Diffuse Rhes neurons are the dominant phenotype and decrease in relative proportion most notably in AD, PiD, and PSP. With increasing Braak stages, there is a negative trend in the quantity of *RASD2* transcripts within two selectively vulnerable excitatory neuron populations, but the groups do not statistically significantly differ from one another (C).

### *RASD2* mRNA levels in selectively vulnerable excitatory neurons of the EC

We analyzed single-nucleus RNA sequencing data from two selectively vulnerable excitatory neuron populations of the EC in a different cohort [26] with three cases at Braak stage 0, four at Braak stage II, and three at Braak stage VI. The mean log10(CPM) transcript levels were computed for each case. Comparisons were made between cases. For each of the two subpopulations, the mean log10(CPM) of *RASD2* transcripts was low (< 0.5). When comparing the mean expression level of *RASD2* mRNA of each case between the three Braak groups, we did not detect significant differences between either group 2 (p=0.286) or group 4 (p=0.404) excitatory neurons of the EC. A negative trend by increased Braak stage was noted, though (Figure 4c).

### TDP-43 mislocalization is not associated with changes in Rhes neuronal phenotype

To examine if abnormal Rhes phenotype also associates with TDP-43 mislocalization, we examined sections from the hippocampal formations of two individuals with frontotemporal lobar degeneration due to TDP-43 Type A immunostained against phospho-TDP and Rhes. Despite the high number of TDP-43 inclusions in neurons observed in both cases, we failed to identify any neurons with Punctiform or Absent Rhes phenotypes (Figure 5).

**Figure 5:**
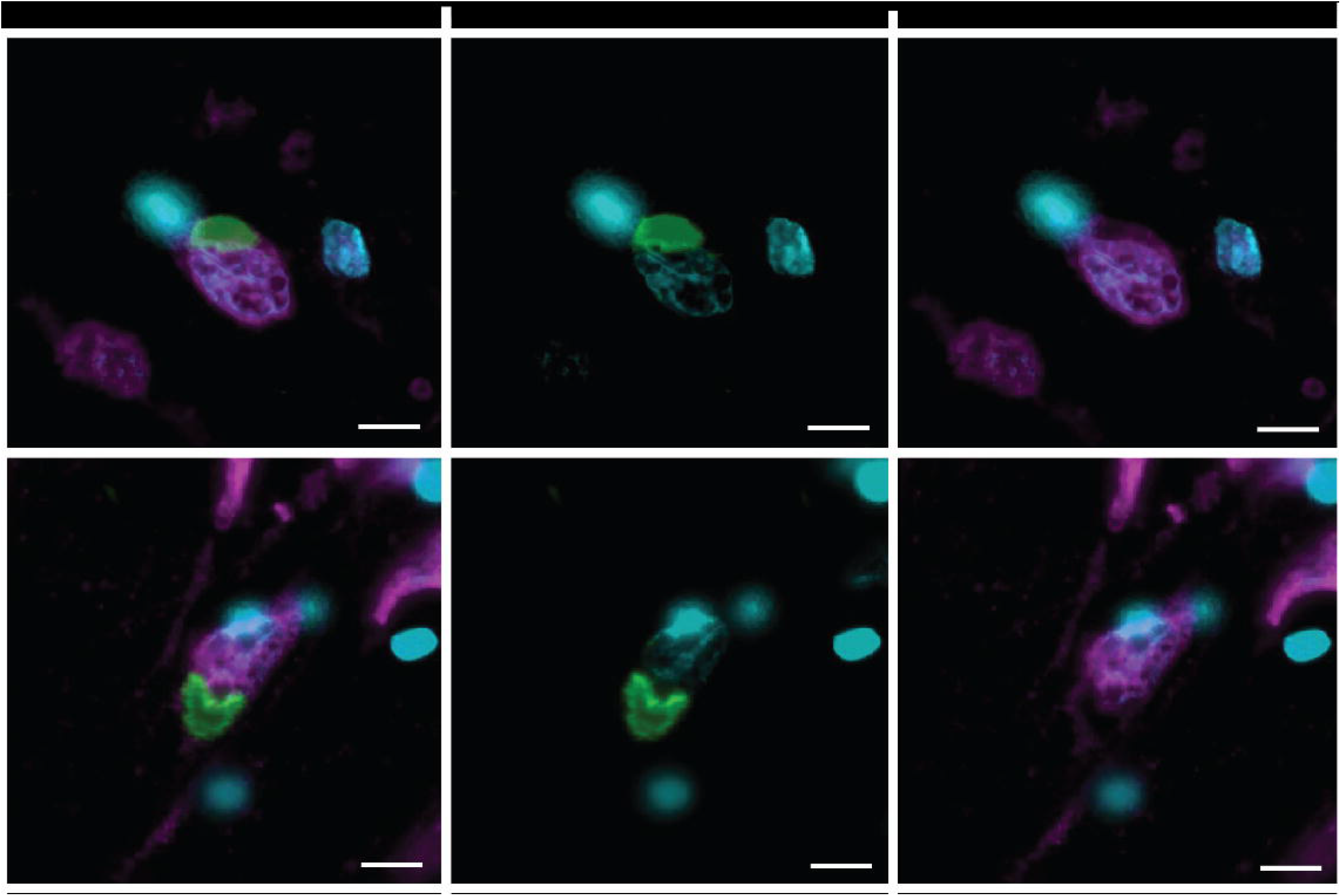
Neurons positive for TDP-43 (green) proteinopathy display a Diffuse Rhes (pink) phenotype. Scale bar represents 10 µm.

### Changes in Rhes subtypes correlate with the presence of intraneuronal tau aggregates

In slides immunostained for Rhes, PHF-1 and NeuN, we counted a total of 5,330 neurons (an average of 296 neurons per case) in the EC. Combining all cases (n=18), only a mean (sd) of 5.1% (0.9%) of diffuse Rhes neurons were PHF-1 positive, 66.0% (35.5%) of punctiform Rhes neurons and 89.7% (23.2%) of absent Rhes neurons were PHF-1 positive (Figure 6a-b, Table 3). The proportion of neurons with PHF-1+ tau inclusions significantly differs between the three neuronal Rhes phenotypes (p < 0.0001). A Kruskal-Wallis test showed that this pattern holds for AD (p = 0.0241), PiD (p = 0.0379), and PSP (p = 0.0347). While not statistically significant in the other tauopathies, the patterns generally followed the same trend.

**Figure 6:**
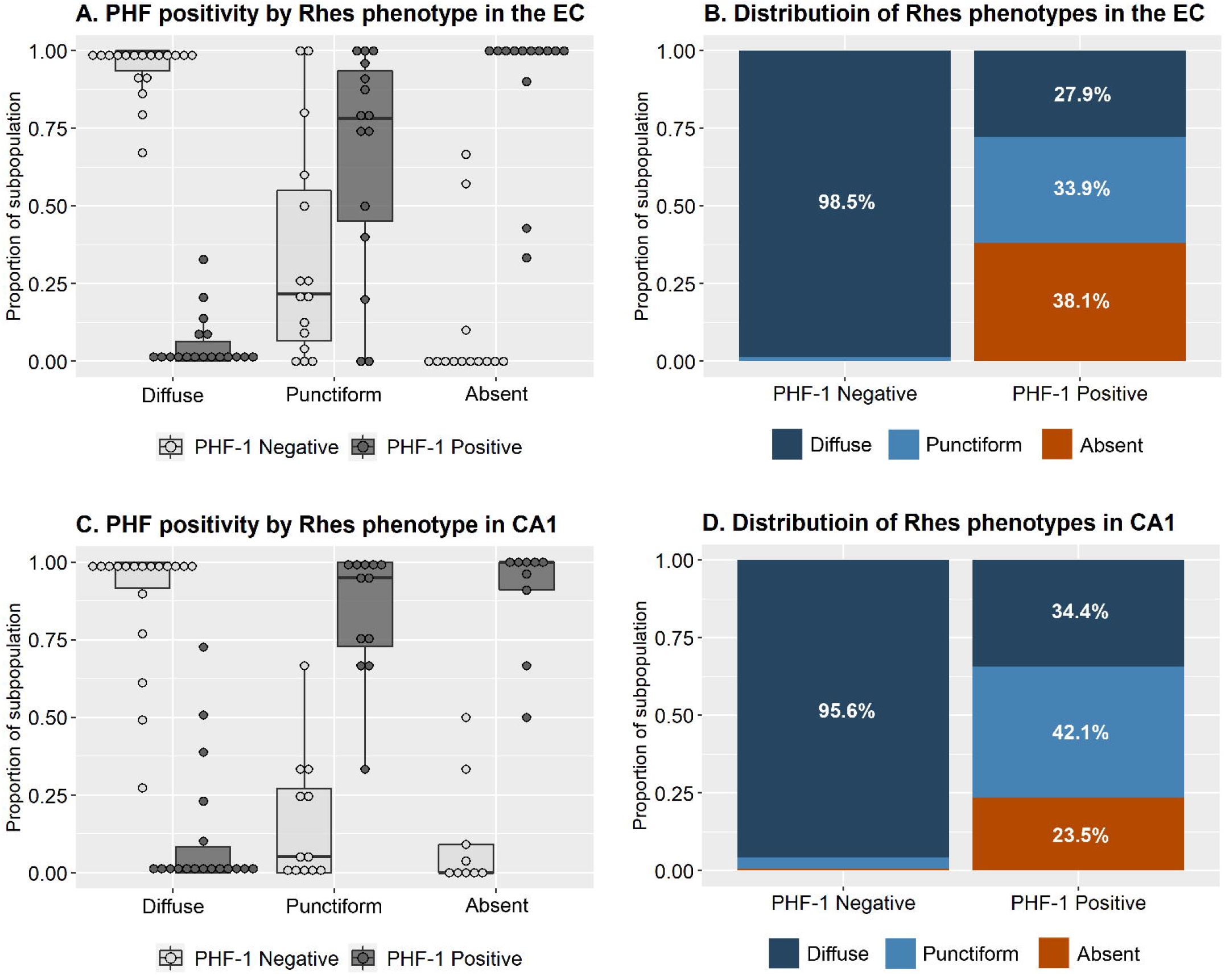
Entorhinal cortex neurons with diffuse Rhes are predominantly PHF-1 negative, while neurons with absent Rhes are predominantly PHF-1 positive (A). While neurons with punctiform Rhes appear to be roughly split between PHF-1 positive and negative neurons in the entorhinal cortex, they make up roughly a third of the PHF-1 positive neurons, indicating a tendency to co-occur with tau inclusions (B). Of the PHF-1 positive neurons in the entorhinal cortex, the majority have either punctiform or absent Rhes. This pattern largely holds for neurons of the cornu ammonis 1 (C & D).

We counted 4,374 neurons (an average of 243 neurons per case) in the CA1 and found similar patterns detected in the EC. Combining all tauopathies (n=18) for the CA1, only a mean (sd) of 11.8% (21.9%) of diffuse Rhes neurons were positive for PHF-1, while 83.8% (20.9%) of punctiform Rhes neurons and 89.3% (18.3%) of absent Rhes neurons were positive for PHF-1 (Figure 6c-d, Table 4). The proportion of neurons with PHF-1+ tau inclusions significantly differs between the three neuronal Rhes phenotypes (p < 0.0001). A Kruskal-Wallis test shows that this same pattern holds for AD (p = 0.0265). While the pattern in other tauopathies is not statistically significant, they do generally follow the same trend.

Given that the pattern was most robustly detected in AD, CA1 from two of the AD cases (59 year-old male and 64 year-old female) were further stained to examine the relationship between Rhes subtypes and other tau post-translational modifications. From those neurons positive for each of phospho-Thr212/Ser214 Tau (AT100), acetyl-Lys274 Tau (1f3c), truncated ΔAsp421 Tau (Tau-C3), and conformationally-changed tau (MC1), the percentage classified as each Rhes phenotype was computed. Overall, the different PTMs featured different abundances of the three Rhes phenotypes, but we failed to identify a tau PTM with >90% correlation with Punctiform and Absent Res phenotype (Table 5). Regardless, for all tau PTMs, diffuse Rhes is the least represented.

## DISCUSSION

As an abbreviation for Ras homolog enriched in striatum, Rhes has been thought to be a striatal-specific protein. Here, we showed that Rhes is expressed throughout the allocortex, confirming other studies suggesting that Rhes expression extends beyond striatal neurons [17, 20]. Not only is Rhes ubiquitously represented in cortical neurons, but it shows a diffuse cytoplasmic distribution in over 99% of these neurons (here defined as Diffuse Rhes type), suggesting that such distribution represents the normal (i.e., healthy) neuronal phenotype. Noticeably, the intraneuronal distribution of Rhes in neurons changes in several sporadic tauopathies, either to a punctiform Rhes positivity in neuronal cell bodies (Punctiform Rhes type) or with an absence of Rhes positivity in neuronal cell bodies (Absent Rhes type). Using cases representing five sporadic tauopathies and healthy controls, we discovered that the proportion of EC and CA1 neurons with the Diffuse Rhes neuronal phenotype decreased in tauopathies.

Neurons with Punctiform and Absent Rhes phenotype were more likely to harbor tau inclusions than neurons with diffuse Rhes. This observation suggested that the population-level decreases in the proportions of neurons with diffuse Rhes in tauopathies associated with either the tau inclusion itself or an upstream process affecting both tau aggregation and Rhes expression. Given the lack of phospho-tau (PHF-1 positive) inclusions in neurons with Diffuse Rhes and its high frequency (>99%) in HC, diffuse Rhes appears to be the normal phenotype. On the other hand, Absent Rhes neurons are predominantly PHF-1 positive, suggesting that abnormal (i.e. diseased) neurons cleared Rhes in the soma. Meanwhile, punctiform Rhes appears to be an intermediate phenotype (Figure 7). Similar patterns were observed for other tau post-translational modifications in AD cases. Punctiform or Absent Rhes were not detected in neurons with TDP-43 Type A presentation. The strong association between abnormal tau inclusions and Punctiform and Absent Rhes phenotypes may explain why CBD and AGD were nearly devoid of changes in Rhes (Figure 4CBD does not significantly target EC and CA1, and AGD is thought to be relatively benign. Furthermore, most tau deposits in AGD are in dendritic buttons rather than the cell body [28, 29]. Notably, the few neurons with tau lesions in CBD and AGD were enriched for Punctiform or Absent Rhes (Tables 3 and 4).

**Figure 7:**
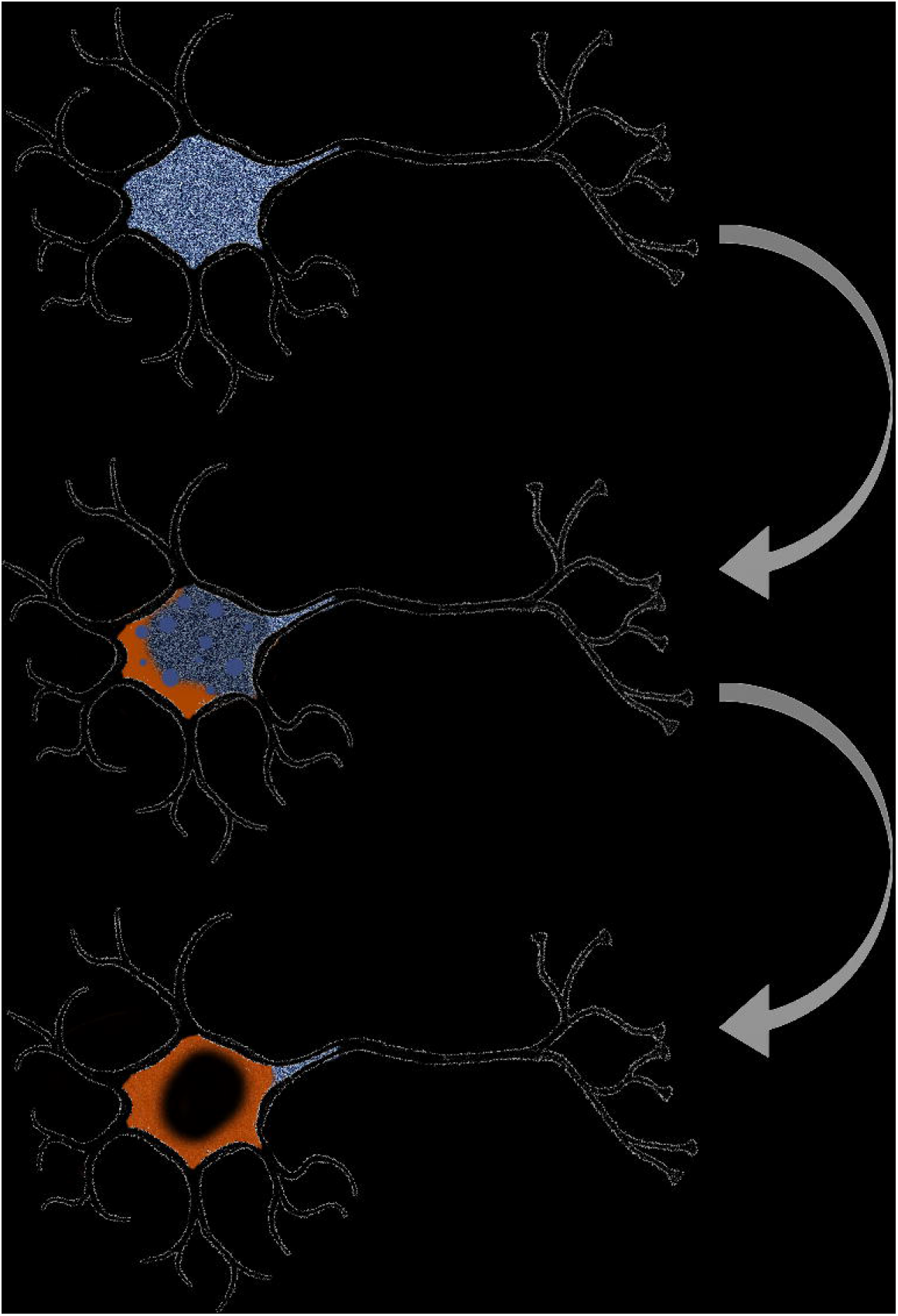
We propose an ordered manner in which the Rhes (blue) phenotypes relate to each other based on the collective evidence from this investigation and others. At baseline, Rhes is expressed throughout the cytoplasm extending into neuronal processes. As neuropathologic changes (orange) emerge, Rhes expression becomes dysregulated and appears punctiform within the cell body. Finally, Rhes is no longer expressed in the cell body of those neurons most affected by neuropathologic changes with some signal persisting in neuronal processes.

Although our findings show robust evidence that Rhes phenotypic changes in neurons are directly associated with tau aggregation, our study design precluded us from determining the direction of the association. We attempted to shed light on this question by interrogating whether RADS2 expression levels decreased first in neuronal subpopulations of the EC deemed more vulnerable to AD than in the other neuronal subpopulations [26] using a dataset with healthy controls and individuals at Braak stage II and IV. We did not detect statistically significant changes in transcript levels of *RASD2* between Braak stages in AD; however, a negative trend began to emerge. Further work with larger RNA-sequencing datasets should be done to understand how expression of *RASD2* changes with disease progression in tauopathies.

This postmortem study in humans together with *in vivo* and *in vitro* work done in models of familial tauopathies [17] favors a causal relationship between Rhes clearance and tau proteinopathic lesions. Given an established role of Rhes in modulating autophagy [16, 17, 19], the high co-incidence of neurons with absent Rhes and PHF-1 positive neurons is not entirely surprising. Hernandez and colleagues [17] demonstrated that pathogenic *MAPT* mutations initially suppress age-related increases in Rhes, congruent with our observations here in sporadic tauopathies. Impaired autophagy is a well-established hallmark of neurodegenerative diseases, including tauopathies [12, 13, 30, 31] and these changes in Rhes distribution may fall within the pathogenic molecular network.

Our results provide a possible explanation for the findings in Hernandez et al [17] demonstrating that lonafarnib was effective in preventing the accumulation of tau proteinopathic lesions in model systems, but not effective in clearing existing tau aggregations. If end-stage neurons feature an absence of Rhes, these neurons would lack the druggable target for lonafarnib to act. In our sample, a notable proportion of PHF-1 positive neurons (23.5-38.1%) had an absent Rhes phenotype, possibly explaining why lonafarnib did not promote clearance of tau aggregations in model systems. Recently, Rhes has been implicated in another potentially relevant mechanism that can impact the pathogenesis of neurodegenerative diseases. Rhes promotes formation of tunneling nanotube□like protrusions, which are thought to facilitate the selective transfer of membrane vesicles and organelles, including lysosomes and endosomes, between cells [32].

Caution should be taken in interpreting the results, as our investigation has limitations. Postmortem studies are cross-sectional in nature. As such, we are unable to assess longitudinal changes in neuronal Rhes phenotypes in tauopathies and specifically how they mechanistically relate to tau. For this reason, our study should be interpreted with others using model systems [17]. Additionally, we have not yet identified sub-cellular structures that colocalize with the puncta observed in punctiform Rhes. Nor have we resolved the pathophysiologic changes that accompany the PHF-1 negative punctiform Rhes neurons. Gaining a better mechanistic understanding of the Rhes neuronal phenotypes in the manner of Figure 6 would be key for precise manipulation of Rhes as a therapeutic target. Finally, although we counted thousands of neurons, they only represented a fraction of the EC and CA1 and do not inform us about other cortical regions also affected by tauopathies. The primary outcomes here were agnostic to the neuropathological diagnosis and focused on the relationship between tau proteinopathic inclusions and changes in neuronal Rhes phenotype in tauopathies as a whole class of diseases which we are sufficiently powered to make conclusions. However, we lacked power to conduct extensive investigation comparing each individual tauopathy.

This study also has several strengths. As with other methods, immunofluorescence may show false positives (off-target signal) despite all measures to avoid it. Here, we validated the finding that Rhes is ubiquitously expressed in cortical neurons using single-nucleus RNA sequencing data from an independent series. We investigated well-characterized postmortem brain tissue of individuals with a range of sporadic tauopathies, making us well-positioned to examine the translatability of results drawn from model systems based on familial tauopathies. One limitation of *in vivo* and *in vitro* tauopathy models is the use of *MAPT* mutations. While also featuring tau aggregations, the specific pathophysiology and etiology of rare familial tauopathies have differences with the more common sporadic tauopathies studied here. Furthermore, it is infeasible to completely model the entire molecular and physiological milieu surrounding neurodegeneration in model systems. As such, it is unclear how generalizable the results of interventions in these model systems are. It is imperative to examine the conclusions of model systems in the context of human cases, as we have done here.

Lonafarnib is a well-tolerated drug for which there is existing clinical experience [33-36]. Here, we show that the expression of a target of lonafarnib, Rhes, is altered in tauopathies, reinforcing the conclusions drawn from model systems. Our findings here, in combination with the mechanistic work done by Hernandez and colleagues [17], lend support to the establishment of a clinical study of lonafarnib for early intervention in tauopathies, including AD.

## Data Availability

Raw data is available upon request

## ACKNOWLEDGMENTS

The authors thank the UCSF Memory and Aging Center patients and their families for their contributions to this work. In particular, we thank those who have donated their brains to the Neurodegenerative Disease Brain Bank. We also thank the staff of the brain bank, without whom this work would not be possible. Additionally, we thank the Grinberg lab staff for their technical and administrative assistance with this work.

Microscopy was done at the Cancer Research Lab Molecular Imaging Center at the University of California, Berkeley. Financial support for the relevant equipment was provided by the UC Berkeley Biological Faculty Research Fund. We thank Feather Ives and Holly Aaron, Ph.D. for their training and assistance.

This study was supported by National Institute on Aging grants K24AG053435, K08AG052648, R01AG062359, F30AG066418 and R56AG057528 as well as National Institute of Neurological Disorders and Stroke grant U54NS100717-04 with additional support from Institutional grants NIH P30 AG062422 and P01AG019724, the Rainwater Charitable Foundation, and the Larry L. Hillblom Foundation.

